# A prospective observational safety study on ChAdOx1 nCoV-19 corona virus vaccine (recombinant) use in healthcare workers- first results from India

**DOI:** 10.1101/2021.04.03.21254823

**Authors:** Upinder Kaur, Bisweswar Ojha, Bhairav Kumar Pathak, Anup Singh, Kiran R Giri, Amit Singh, Agniva Das, Anamika Misra, Ashish Kumar Yadav, Sankha Shubhra Chakrabarti, Sangeeta Kansal

**Author notes:** Corresponding author: Prof Sangeeta Kansal, Department of Community Medicine, Institute of Medical Sciences, Banaras Hindu University, Varanasi, UP, India, PIN-221005. Co-corresponding author: Dr SS Chakrabarti, Department of Geriatric Medicine, Institute of Medical Sciences, Banaras Hindu University, Varanasi, UP, India, PIN-221005.

## Abstract

Vaccines are an important public health measure for tiding over the COVID-19 pandemic. Several vaccines have been approved in different countries for emergency use. In India, two vaccines have been currently approved-COVISHIELD (Serum Institute of India (SII)) which is a recombinant simian adenovirus-based vaccine and COVAXIN (Bharat Biotech) which is an inactivated SARS-CoV-2 vaccine. Our current study provides the first post approval safety data on ChAdOx1 nCoV-19 corona virus vaccine (recombinant) use in healthcare workers in northern India (n=804). Around one half of vaccine recipients developed adverse events at any time post vaccination with majority of reactions being mild to moderate in severity. AEFIs were seen in 40% participants after first dose and around 16% participants after second dose. This observed reactogenicity is much less compared to 60-88% reactogenicity rate observed with Oxford-AstraZeneca’s ChAdOx1 vaccine in the UK based population. Individually, fever, injection site pain and headache were the commonly observed AEFIs. Overall, the frequency of systemic events of severity grade 3 was only 0.5% and is much less than the reported rates for other recombinant adenoviral vaccines. The rate of serious AEFIs in our study was only 0.1% (n=1). There was a possibility of this AEFI being an immunization stress related response. No deaths were reported in the vaccine recipients in our study during the study period. Reactogenicity rate was observed to decrease with age and was higher in females. On the basis of interim findings of this safety study, it may be interpreted that the ChAdOx1 nCoV-19 corona virus vaccine (recombinant) (COVISHIELD, Serum Institute of India) carries a good safety profile overall.

## 1. Introduction

COVID-19 is an infectious respiratory illness caused by a novel beta corona virus, SARS-CoV-2. The course of COVID-19 disease can be unpredictable and mortality as high as 26% has been observed in the elderly population and those with co-morbidities.(1) Deaths due to COVID-19 are often because of respiratory failure, septic shock, disseminated intravascular coagulation, and sometimes myocardial injury.(2) The treatment of COVID-19 at present relies on supportive therapies such as prophylactic antibiotics, oxygen supplementation and parenteral steroids. In the absence of definitive anti-SARS-CoV-2 therapy, immunization against viral disease or at least against severe form of illness may offer an attractive means of curtailing the epidemic. This unmet need spurted the development of vaccines which are being manufactured using pre-existing and novel platforms and are in various preclinical and clinical phases. Some of these vaccines such as Moderna’s mRNA-1273, Pfizer’s mRNA based BNT162b2, Oxford university-Astra Zeneca’s vaccine based on the simian adenovirus have been given emergency use authorization status in various countries. In India, COVISHIELD (Serum Institute of India (SII)) and COVAXIN (Bharat Biotech) have been approved for emergency use. COVISHIELD is based on a replication-deficient simian adenoviral vector coding the whole length spike glycoprotein (S) of SARS-CoV-2 while COVAXIN is based on inactivated SARS-CoV-2 platform. The vaccines have been rolled out all over India and are being administered to all individuals ≥ 18 years of age other than those with a history of allergy to one of its components. The first phase of vaccination was directed towards health care workers and front-line workers (police, sanitary workers etc) who are at increased risk of acquiring COVID-19, and who consented to receiving the vaccines. However, the type of vaccine allocated for a particular center (COVISHIELD or COVAXIN) is at the discretion of the government and based on availability status and logistic concerns. Both the vaccines are being provided free of cost by the government of India, through state government health systems and utilising an elaborate and well-designed micro plan of vaccinating every front-line worker. Pre-approval COVID-19 vaccine trials have been done largely in healthy population under controlled settings, have limited inclusion of diverse ethnicities and are limited by short duration of follow up with merging of various phases of clinical trials. Such studies therefore may not detect all safety-related issues that arise when vaccines are intended for marketing in general population. The main objective of this observational study is to carry out a detailed long term safety analysis of COVISHIELD use in the Indian population. Here we present the first interim safety analysis of use of COVISHIELD in health care workers in three vaccination centres in the city of Varanasi (Uttar Pradesh) in north India. COVISHIELD was the designated vaccine for these centres and hence the focus of our study.

## 2. Materials and Methods

### 2.1. Study design and setting

Ours is a continuing prospective observational study which started from 5^th^ Feb 2021 and is expected to be continued till May 2022 with at least one year follow up of all the recipients enrolled. The study is being conducted at three sites in Varanasi: Sir Sunderlal hospital which is one of the largest tertiary care teaching and research hospitals of north India, SVM hospital which is a district hospital, and urban community health centre (UCHC), Durgakund. Here we report the first results of a subset of participants who have been followed up for at least seven days post second dose of vaccination.

### 2.2. Study participants

All the individuals who received vaccines, in the above-mentioned centres, and who provided consent to participate were enrolled in the study. In the current analysis, all enrolled participants are healthcare workers. The study involves follow up of the enrolled individuals for at least one year.

### 2.3. Safety analysis

Adverse events following immunization (AEFIs) were recorded at prespecified intervals and the following detailed data for safety analysis were extracted.

- Incidence of AEFIs
- Type and pattern of AEFIs
- Distribution of AEFIs with respect to age and gender
- Outcomes of AEFIs
- Seriousness of AEFI as per WHO definition
- Severity of AEFIs for local AEs (adverse events), systemic AEs, and vital signs. These were recorded as per FDA severity grading scales of individual AEFIs and as per modified Hartwig’s severity scale of ADR (adverse drug reaction) for AEFIs not mentioned in the FDA guidance document.
- Causality assessment of AEFIs by WHO Scale
- AEFIs resulting in hospitalization.
- Any vaccine-disease interaction resulting in AEFI
- Any vaccine-drug interaction resulting in AEFI

### 2.4. Vaccination procedure and enrolment in study

As per government policy, COVISHIELD is being administered to health care workers and frontline workers, in the centres of the current study. The vaccine is administered in a dose of 0.5 mL in a two-dose schedule, with the doses given at interval of 4-6 weeks (now revised to 8-12 weeks), intramuscularly in the deltoid. Each mL of the dose administered contains 5 × 10^10^ simian adeno-viral particles produced in genetically modified human embryonic kidney (HEK) 293 cells. All recipients are routinely monitored at study sites for 30 minutes post vaccine administration, as a part of standard operating procedure for vaccination. All those participants who gave consent to participate in our study were enrolled and are being contacted on phone after 24 hours of vaccination, at day 7, day 14, day 28 and thereafter monthly for a total period of one year. A support phone number is provided to each participant to contact for reporting, at times of emergency or in case of any doubts. For safety analysis, individuals are specifically questioned about local site symptoms such as pain, erythema, swelling, tenderness, and degree of limitation of physical activity. They are also questioned about systemic symptoms such as fever, fatigability, myalgia, arthralgia, headache, nausea, vomiting, diarrhoea, rash, chest tightness and dyspnoea. Biochemical tests are not done routinely in all the vaccine recipients but are planned in case of persistence or severe form of AEFIs. Individuals are informed about the clinical features of COVID-19 and are instructed for RT-PCR based nasal or oropharyngeal swab test for SARS-CoV-2 in any event of developing COVID-19 like symptoms. However, since the study is focused on safety analysis only, testing was not compulsory and only at the discretion of the participants.

### 2.5. Ethical permission

The study started after obtaining permission from the Ethics Committee of the Institute of Medical Sciences, Banaras Hindu University, and written informed consent was taken from all the participants.

### 2.6. Data sources/ measurement

Data pertaining to demography, medical history including history of SARS-CoV-2 positivity at any time in the past, existing co-morbidities, concurrent drug history and history of allergy to any known stimuli are recorded in a pre-designed case report form. Information regarding development of AEFIs, severity of AEFIs, interventions required for management of AEFIs, outcomes of AEFIs, time to complete recovery, and causality of AEFIs is also collected. Causality assessment of serious AEFIs (WHO classification) and AEFIs with FDA grade 3 or more was done by the investigators of the study.

### 2.7. Sample size

So far, the trials analysing the safety and reactogenicity of COVID-19 vaccines have reflected inconclusive evidence on the rates of occurrence of adverse events of clinical significance. Clinically significant AEFIs have been seen to occur in 1-20% of COVID-19 vaccine recipients. In view of lack of India-specific data and assuming an average rate of occurrence of clinically significant AEFI to be 10% and margin of error of 2.5%, the expected sample size for this study was calculated to be 576. After clinical and feasibility considerations, the authors decided to include at least 1400 vaccine recipients for detailed analysis. Considering a drop-out rate of 15%, it was planned to enroll at least 1650 individuals.

### 2.8. Statistical analysis

Results were recorded as percentages as well as frequencies for data such as incidence, type, severity, and outcomes of AEFIs. Independent t test was used to compare the quantitative variables such as age, between the group developing AEFIs and that without AEFIs. Chi square test was applied for dichotomous variables such as gender, presence of co-morbidities and co-medications to find association between various factors and development of AEFIs.

## 3. Results

Figure 1. represents the enrolment of vaccinated individuals in the present study. A total of 1666 individuals were screened of whom 16 refused to participate in the study. Of the included 1650 participants, 846 and 804 participants respectively were visiting the centres for their first and second dose of vaccine. A significant percentage of the subset who were enrolled at the time of receiving the first dose is yet to receive their second dose, as timing of second dose has been changed from 4 weeks to 8-12 weeks, after initiation of our study. These individuals shall be included in the full analysis of study which is planned early next year. For the other 804 individuals who were enrolled in the study while receiving the second dose of vaccine, detailed enquiry was made about any AEFIs during their first dose of vaccine as per protocol described in methods section. They were subsequently followed up after their second dose. Total period of follow-up was calculated starting from their day of receiving first dose and up to 12^th^ March 2021. Median (Q1,Q3) follow up period was 42 (36,43) days. Of these 804, two participants were considered ineligible for the second dose of vaccine by the vaccination programme authorities because of possible AEFI concerns. The investigators of the current study had no role in determining this ineligibility for vaccination. The baseline characteristics of the study participants are mentioned in **Table 1**.

**Figure 1:**
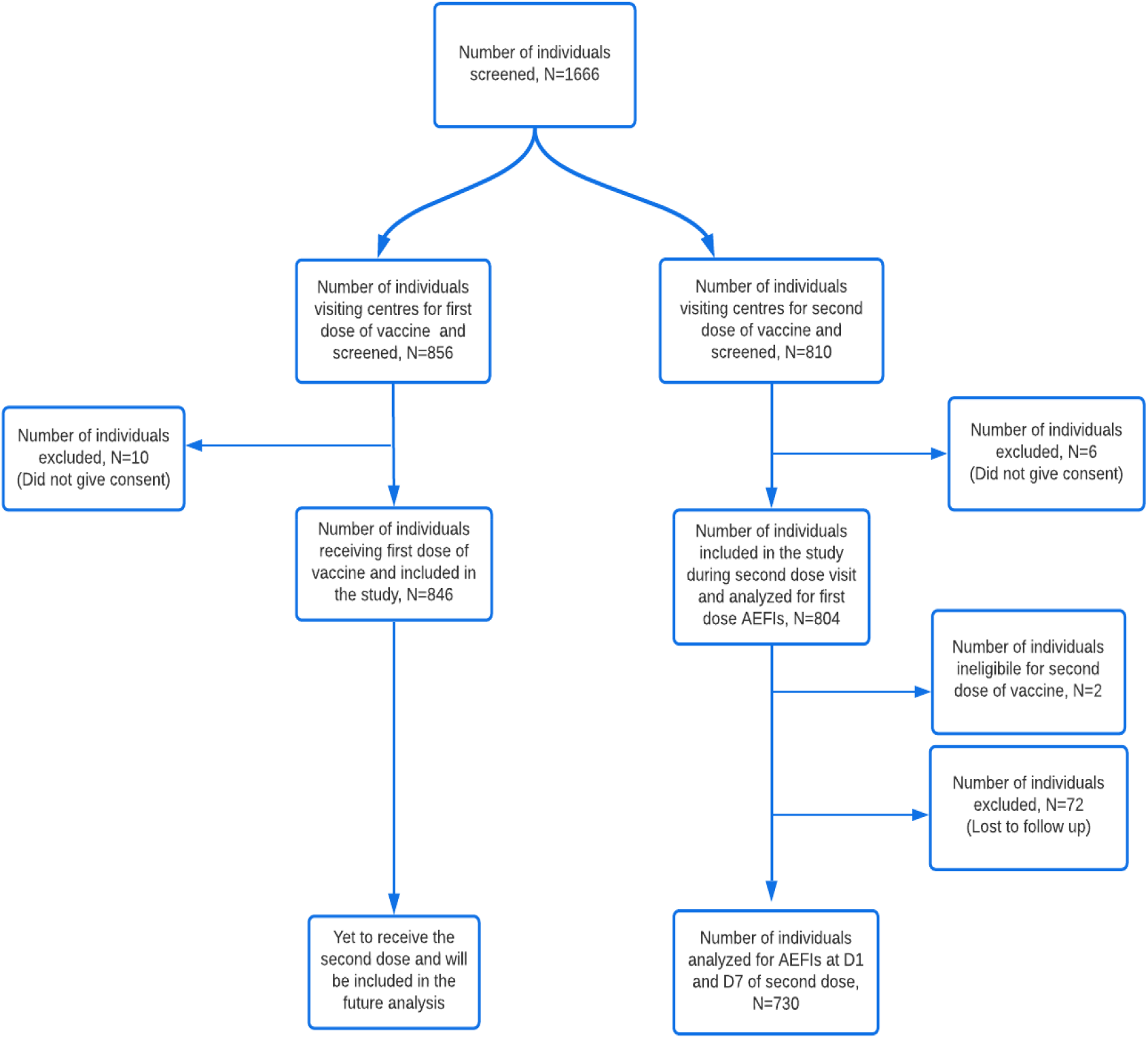
Flowchart showing enrollment of participants in safety study.

### 3.1. AEFIs after first dose of vaccine

Of the 804 vaccine recipients, AEFIs were reported in 321, giving an AEFI incidence rate of 40%. Systemic AEFIs with or without local (injection site) involvement was seen in around 31% participants and only local site involvement was observed in 9% individuals. Among systemic AEFIs, fever, malaise and headache were the commonly reported AEFIs, seen respectively in 15.2%, 8.7% and 5.8% individuals. Severity wise, 70.4% AEFIs were classified as ‘mild’ and 28.7% were of ‘moderate’ category. Two AEFIs (0.62%) were of grade 3 severity and one AEFI (0.3%) was ‘serious’ and led to hospitalization. The single serious AEFI was classified as ‘possible’ on causality assessment and with suspicion of immunization stress related response (ISRR). Median time of complete recovery from AEFIs was 1 day. 91 participants with AEFIs needed interventions of which paracetamol was used in 82 cases, anti-histaminics in 7 cases, and tramadol in 2 cases.

### 3.2. AEFIs within 30 minutes of second dose

Out of total 802 participants, AEFIs were observed in seven individuals (0.9%). Three recipients developed systemic AEFIs while only local involvement was seen in four participants. All seven AEFIs were of ‘mild’ severity. Median time to complete recovery was 2 days.

### 3.3. AEFIs within 24 hours and till day 7 post-second dose

After excluding 72 individuals who were lost to follow up, a total of 730 individuals were included for analysis of AEFIs occurring within 24 hours, and between 24 hours and 7 days of vaccination, but not within 30 mins post vaccination. Of these, 93 vaccine recipients (12.73%) developed AEFIs within 24 hours and 22 (3%) developed AEFIs after day 1 and till day 7 post-vaccination, respectively. AEFIs were thus observed in a total of 115 recipients (15.7%) till day seven. Systemic involvement with or without local site reaction was seen in 99 (13.6%) and only local involvement was seen in 16 (2.2%). Severity wise, two recipients developed AEFIs of grade 3 severity (causality assessment-1 probable, 1 possible). No serious AEFIs or deaths were reported in this subset of 730 recipients. On performing causality assessment, AEFIs belonged to probable, possible, and unclassifiable categories in 104 (90.4%), 4 (3.5%), and 7 (6%) cases, respectively. Of those developing AEFIs, 41 (35.6%) recipients required interventions. Paracetamol was required in 36, antibiotics in 5, proton pump inhibitors in 3, anti-histaminics in 2, anti-emetics and intravenous fluids in 1. Median time to complete recovery was 2 days for AEFIs developing within 24 hours of vaccination, as well as for AEFIs developing between 24 hours and 7 days of vaccination. Common AEFIs following second dose of vaccine were fever, injection site pain, and headache.

## 4. Discussion

The results of this interim analysis show that ChADOx1 vaccine (Serum Institute of India) has a generally favourable safety profile. Around one half of vaccine recipients developed adverse events at any time post vaccination with majority of reactions being mild to moderate in severity. AEFIs were seen in 40% participants after first dose and around 16% participants after second dose. This observed reactogenicity is much less compared to 60-88% reactogenicity observed in phase 1 and phase 2/3 clinical trials of Oxford-AstraZeneca’s ChAdOx1vaccine in the UK based population (AZD1222).(3,4) Systemic involvement with or without local site involvement was seen in nearly one third of vaccine recipients after first dose and in 13.6% vaccine recipients after second dose. Fever, injection site pain and headache were the commonly observed AEFIs. Fever occurred in 15% individuals after first dose and in 3-4% participants after second dose. Frequency of other events such as malaise, and headache remained low at the rate of 3-4%. Previously, these events have been shown to occur in 30-70% UK based recipients of Oxford-AstraZeneca’s ChAdOx1 vaccine, 40-50% Chinese recipients of recombinant Ad5 based vaccine manufactured by CanSino Biologics and in 20-40% US and Belgium based individuals receiving recombinant Ad26 based vaccine of Janssen Pharmaceuticals.(3–6) The frequency of AEFIs decreased with age, with around 27% lower risk of development of AEFIs in participants of ≥ 40 years age compared to those in the 18–39 years age group. That increasing age is associated with lesser risk of AEFIs is in concordance with the published clinical trials analysing various viral vector-based vaccines.(3,5,7) No significant association of AEFI risk was seen with lab confirmed diagnosis of COVID-19 in past. Likewise, co-morbidities such as diabetes mellitus and asthma or COPD did not have any statistically significant association with AEFIs. The findings should be interpreted with caution as majority of vaccine recipients enrolled in the study were healthy individuals and co-morbidities were present in only around 10% of individuals.

Severity wise, four participants developed AEFIs of grade 3 severity assessed by the Food and Drug Administration (FDA) toxicity grading scale. Two of these vaccine recipients developed symptoms after second dose and two after the first dose. One of the latter two was considered ineligible for second dose by the vaccination programme authorities. Three grade 3 events were rated as ‘probable’ and one as ‘unclassifiable’ by the investigators using the WHO scale of causality assessment. One patient developed serious (WHO) AEFI leading to emergency visit followed by in-patient ward admission. The patient was discharged within 2 days with a final diagnosis of COVID-19 vaccination reaction. A possibility of immunization stress related response (ISRR) existed in the case and the reaction was scored under ‘possible’ category by the investigators. The lady refused second dose of vaccination and was also considered ineligible for second dose by the vaccination programme authorities. Three of the five patients developing AEFIs of ≥ 3 grade were females.

Overall, the frequency of systemic events of severity grade 3 was 0.5% and is much less than the reported rates of 9-20% with rAd5 and rAd26 based vaccines.(5,7) The interim analysis of clinical trials investigating ChAdOx1 in UK, South Africa and Brazil showed a 0.7% rate of occurrence of serious adverse events. The corresponding rate in our study was 0.1% (1/730). No deaths were reported in the vaccine recipients in our study during the study period.

Low reactogenicity rates with COVISHIELD (Serum Institute of India) compared to Oxford-AstraZeneca’s ChAdOx1 vaccine and other adenovirus-based vaccines can be explained to a certain extent by pre-existing immunity against human and chimpanzee adenoviruses in the Indian population by virtue of exposure to such viruses in the past. Human adenoviruses, known to cause common cold in humans are widely prevalent in developing countries. Neutralizing antibodies (nAbs) against human Ad5 have been observed in 100% healthy Indians with medium to high titre nAbs seen in 50% and very high titre in around 30% samples.(8) On the other hand, frequencies, and mean titre of such nAbs are low in the US population.(9) Neutralizing antibodies against chimpanzee adeno viruses are less common but seen in <15% Americans, Chinese and Europeans.(10) Though pre-existing humoral immunity against human adenoviruses is unlikely to cross react with chimpanzee adeno virus ChADOx1, T cells mounted against human adeno viruses are known to cross react with some viruses such as ChAd6 and ChAd7.(10) Humans being in a close phylogenetic relationship with chimpanzees, a possibility of cross reactivity between human and chimpanzee adeno viruses exists. Zhu et al in the phase 2 study on the recombinant Ad5 based vaccine demonstrated that pre-existing neutralising antibodies against human adenovirus 5 might be responsible for low reactogenicity in the elderly compared to the young, which is also a finding of our study.(7) Cross reactivity to other viruses has also been hypothesized by the authors of the current study as a reason for country specific variations in COVID-19 outcomes.(11)

## 5. Limitations of study

By the time the study received permission from Institute Ethics Committee, a subset of participants had received their first dose of vaccination. Information regarding AEFIs throughout 4-6 weeks following the first dose was collected from this subset at the time of their second dose of vaccination. A possibility of recall bias and uncertainty regarding some of the parameters such as time to full recovery exists. The authors however do not think that a gap of 4-6 weeks would have significant clinical bearing on the overall analysis. Further attempt was made to verify the AEFIs from caregivers or close family members and to verify all serious AEFIs occurring during this time from any existing medical records of the recipients. Being an unblinded study, the possibility of observer bias, however, cannot be ruled out. Similarly, as blood investigations were not routinely performed, some AEFIs may have been missed.

## 6. Conclusion

On the basis of interim findings of this safety study, it may be interpreted that the ChAdOx1 nCoV-19 corona virus vaccine (recombinant) (COVISHIELD, Serum Institute of India) carries a good safety profile overall. Reactogenicity decreased with increasing age. In line with the published international evidence on ChAdOx1 nCoV-19 corona virus vaccine, majority of AEFIs are mild to moderate AEFIs and mostly self-resolving. Larger double blind randomized clinical trials of longer duration will give a more appropriate idea of overall safety and efficacy of COVID-19 vaccines.

## Data Availability

Data will be made available to individual researchers if requested.

