## Supplementary material for "A prospective observational safety study on ChAdOx1 nCoV-19 corona virus vaccine (recombinant) use in healthcare workers- first results from India": Table 1

| Vaccinated individuals enrolled (N) | 804 |
| --- | --- |
| Age (years); Mean (± SD) | 38.44 (± 11.47) |
| Gender (Male/Female) | 573/231 |
| Body mass index (kg/m^2^); Mean (± SD) | 24.68 (± 3.68) |
| History of laboratory confirmed COVID-19 at any time before vaccination; N (%) | 56 (7) |
| Blood Group | N (%) |
| B^+^ | 252 (31.3) |
| O^+^ | 225 (28) |
| A^+^ | 132 (16.4) |
| AB^+^ | 66 (8.2) |
| B^-^ | 13 (1.6) |
| O^-^ | 7 (0.9) |
| AB^-^ | 4 (0.5) |
| A^-^ | 4 (0.5) |
| No details | 101 (12.5) |
| Individuals with diabetes mellitus; N (%)  On antidiabetic drugs; N (%) | 66 (8.2)  51 (6.3) |
| Individuals with hypertension; N (%)  On anti-hypertensive drugs; N (%) | 73 (9)  71 (8.8) |
| Individuals with hypothyroidism; N (%)  On thyroxine; N (%) | 28 (3.5)  27 (3.3) |
| Individuals with asthma or COPD; N (%)  On inhaled beta agonists; N (%)  On inhaled steroids; N (%) | 10 (1.2)  5 (0.6)  4 (0.5) |
| Individuals with coronary artery disease; N (%)  On antiplatelet drugs; N (%)  On statins; N (%) | 5 (0.6)  4 (0.5)  3 (0.4) |
| Individuals with self-described allergy to any agent (environmental; household; medications etc); N (%) | 51 (6.3) |
| Individuals with past history of or active tuberculosis; N (%)  On anti-tubercular therapy; N (%) | 4 (0.5)  3 (0.4) |
| Individuals with epilepsy; N (%)  On antiepileptic drugs; N (%) | 2 (0.25)  2 (0.25) |
| Individuals with skin diseases; N (%) | 3 (0.4) |
| Individuals with rheumatoid arthritis; N (%) | 3 (0.4) |
| Individuals currently receiving other drugs | N (%) |
| Non-steroidal anti-inflammatory drugs  Antibiotics  Anticoagulants | 3 (0.4)  1 (0.1)  1 (0.1) |

**Table 1: Baseline characteristics of study participants**

***Abbreviations:*** SD, standard deviation; COPD, chronic obstructive pulmonary disease. All percentages in brackets are out of total enrolled vaccinees (N=804).
